# Performance characteristics of the VIDAS^®^ SARS-COV-2 IgM and IgG serological assays

**DOI:** 10.1101/2020.09.28.20196030

**Authors:** Nathalie Renard, Soizic Daniel, Nadège Cayet, Matthieu Pecquet, Frédérique Raymond, Sylvie Pons, Julien Lupo, Carole Tourneur, Catherine Pretis, Guillaume Gerez, Patrick Blasco, Maxime Combe, Imen Canova, Mylène Lesénéchal, Franck Berthier

**Author notes:** Nathalie Renard, Mylène Lesénéchal.

## Abstract

The COVID-19 pandemic, caused by the new severe acute respiratory syndrome coronavirus 2 (SARS-CoV-2), continues to spread worldwide. Serological testing for SARS-CoV-2-specific antibodies plays an important role in understanding and controlling the pandemics, notably through epidemiological surveillance. Well validated and highly specific SARS-CoV-2 serological assays are urgently needed. We describe here the analytical and clinical performance of VIDAS^®^ SARS-CoV-2 IgM and VIDAS^®^ SARS-CoV-2 IgG, two CE-marked, EUA-authorized, automated, qualitative assays for the detection of SARS-CoV-2-specific IgM and IgG, respectively. Both assays showed high within-run and within-laboratory precision (coefficients of variation < 11.0%) and very low cross-reactivity towards sera of patients with a past common coronavirus or respiratory virus infection. Clinical specificity determined on up to 989 pre-pandemic healthy donors was ≥ 99% with a narrow 95% confidence interval for both IgM and IgG assays. Clinical sensitivity was determined on up to 232 samples from 130 RT-PCR-confirmed SARS-CoV-2 patients. The positive percent agreement (PPA) with SARS-CoV-2 PCR reached 100% at ≥ 16 days (VIDAS^®^ SARS-CoV-2 IgM) and ≥ 32 days (VIDAS^®^ SARS-CoV-2 IgG) of symptom onset. Combined IgM/IgG test results improved the PPA compared to each test alone. SARS-CoV-2 IgG seroconversion followed closely that of SARS-CoV-2 IgM and remained stable over time, while SARS-CoV-2 IgM levels rapidly declined. Interestingly, SARS-CoV-2-specific IgM and IgG responses were significantly higher in COVID-19 hospitalized vs. non-hospitalized patients. Altogether, the VIDAS^®^ SARS-CoV-2 IgM and IgG assays are highly specific and sensitive serological tests suitable for the reliable detection of past acute SARS-CoV-2 infections.

## INTRODUCTION

Coronavirus disease 19 (COVID-19) is an infectious disease caused by the newly discovered severe acute respiratory syndrome coronavirus 2 (SARS-CoV-2) (1, 2). Within three months of its emergence in China in December 2019, COVID-19 has been declared a global pandemic by the World Health Organisation (WHO). As of October 29, 2020, nearly 45 million COVID-19 cases and 1.2 million deaths have been reported worldwide (3–5). Accurate diagnosis is essential in managing the pandemic, not only to identify, isolate and treat affected patients, but also to characterize the epidemiology of virus transmission and develop national and international surveillance programs. WHO recommends molecular testing of SARS-CoV-2 nucleic acids for acute-phase diagnosis of suspected cases (6–8). Several nucleic acid amplification tests (NAATs), mostly based on quantitative reverse transcriptase polymerase chain reaction (RT-qPCR), have received the Conformité Européenne (CE) mark and have been approved by the United States Food and Drug Administration (FDA) under emergency use authorization (EUA) (9–11). On the other hand, serological testing for SARS-CoV-2-specific antibodies, especially immunoglobulin M (IgM) and immunoglobulin G (IgG), is not recommended as the primary method for the diagnosis of acute cases. It plays however an essential role in the diagnosis of past SARS-CoV-2 infection and in ongoing immunological and epidemiological surveillance. Serological testing might also complement molecular testing to confirm suspected cases not detected by molecular assays, either due to late (> 7 days after infection) or improper sample collection. Finally, serology screening may allow the identification of convalescent plasma donors for use as potential therapy against COVID-19 (9–18).

SARS-CoV-2 serological testing is facing several challenges. Among them, sensitivity and specificity should be well defined for the target population and validated at different post-infection time windows. Specificity is particularly critical in the current pandemic phase, as seroprevalence in the population is still low. In such low-incidence settings, a specificity > 99% and a narrow 95% confidence interval (95% CI) are required to ensure a high positive predictive value (PPV) (11, 19, 20). Accordingly, the antigens used to design serology tests should be properly selected and cross-reactivity with antibodies directed against other antigens, including from other coronaviruses, should be verified. A huge number of serology assays have been developed and marketed in the last few months, of which 56 received FDA’s EUA (as of October 29, 2020) (21, 22). Clinical performance data of commercial tests are still scant, and examples of poorly performing tests have even been reported (9). Therefore, there is an urgent need for well validated and performant serology tests, notably demonstrating very high specificity.

We describe here the analytical and clinical performance of VIDAS^®^ SARS-CoV-2 IgM and VIDAS^®^ SARS-CoV-2 IgG, two CE-marked and EUA-authorized automated qualitative assays for the detection of SARS-CoV-2-specific IgM and IgG, respectively, in serum or plasma. Kinetics of SARS-CoV-2-specific IgM and IgG seroconversion using the VIDAS^®^ SARS-CoV-2 IgM and VIDAS^®^ SARS-CoV-2 IgG assays were also compared in hospitalized and non-hospitalized COVID-19 patients.

## MATERIALS AND METHODS

### Patients and samples

SARS-CoV-2-positive samples were collected after approval by the Ethics Committee RCB 2020-A00932-37. Informed consent was obtained in accordance with the local regulations. Pre-pandemic samples (from healthy subjects and from donors with other medical conditions) were collected in accordance with the Declaration of Helsinki, as revised in 2013. Collected sera and plasma were stored frozen (< -20°C) until further testing.

Serum from up to 989 healthy pre-pandemic adult donors collected before September 2019 at two geographical sites (Etablissement Français du Sang (EFS), France; Clinilabs, Inc., United States) were used to determine the assay specificity of the VIDAS^®^ SARS-CoV-2 IgM, IgG and combined IgM/IgG test (defined as negative if both VIDAS^®^ SARS-CoV-2 IgM and IgG assays are negative).

For the evaluation of the positive percent agreement (PPA), 405 serum or plasma samples from 142 symptomatic patients (60 hospitalized, 61 non-hospitalized, 21 of unknown hospitalization status) diagnosed with COVID-19 and confirmed positive for SARS-CoV-2 by molecular testing (cobas^®^ SARS-CoV-2, Roche 09175431190 or Real-time RT-PCR assays for the detection of SARS-CoV-2 Institut Pasteur, Paris (23), performed at the collection site) (Fig. 1) were collected at three local hospitals (Centre Hospitalier Saint Joseph Saint Luc, Lyon, France; Centre de Ressources Biologiques [CRB] des Hospices Civils de Lyon, CRB Nord and CRB Sud, Lyon, France) between March 31 and June 2, 2020. Samples were tested with the VIDAS^®^ SARS-CoV-2 IgM and IgG assays, and paired measurements were considered for the combined IgM/IgG test results (defined as positive if at least one of the VIDAS^®^ SARS-CoV-2 IgM and/or IgG assays is positive) (Fig. 1). The PPA was evaluated according to weekly time frames (0-7, 8-15, 16-23, 24-31, ≥ 32 days) relative to the time from RT-PCR positive result and from symptom onset, when documented (Fig. 1).

**Figure 1.**
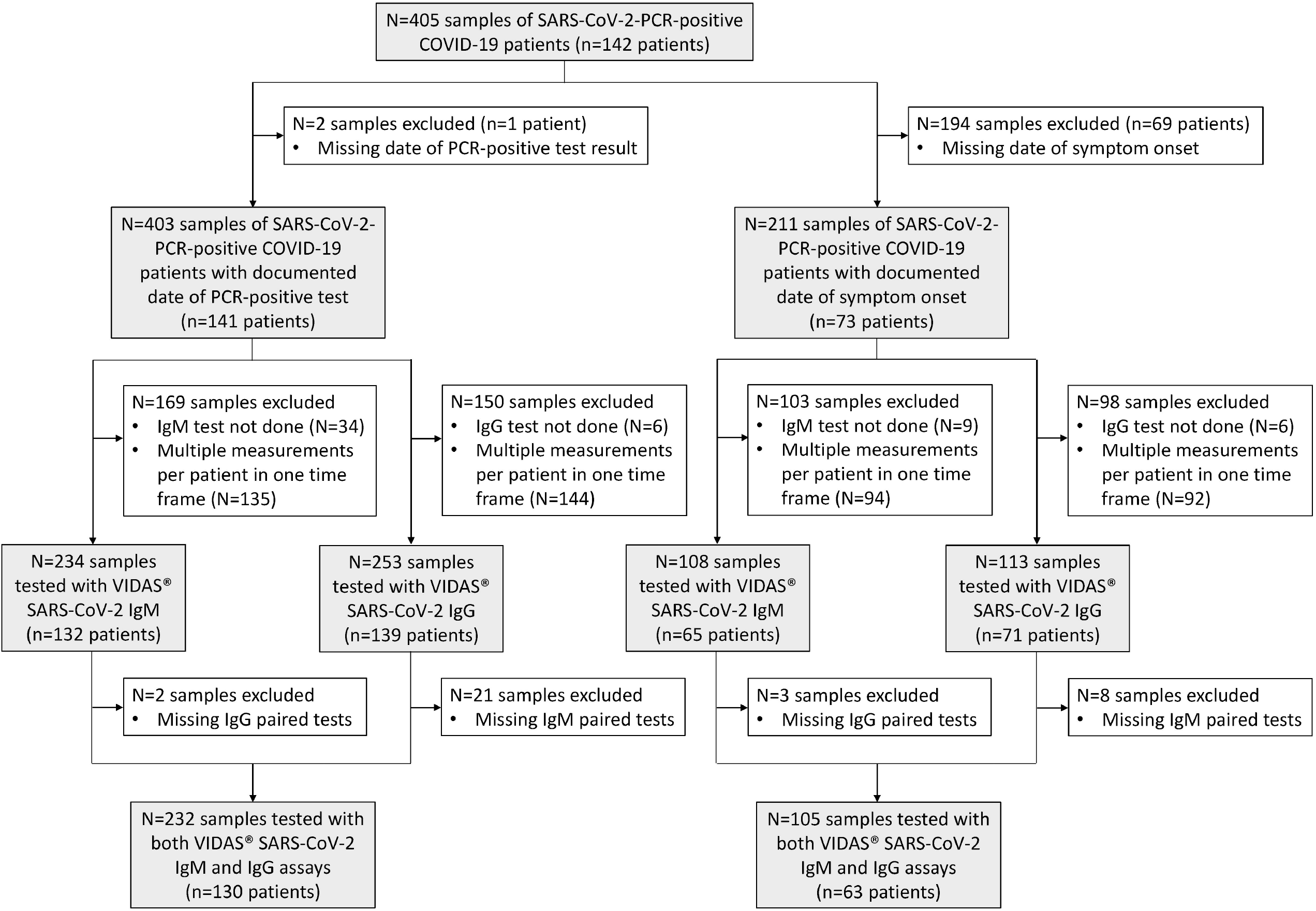
Study flow diagram. Description of SARS-CoV-2-positive samples used to determine the positive percent agreement relative to the time of RT-PCR-positive test result and to the time of symptom onset. The number of and reason for sample exclusion are indicated in the white boxes. Included samples are indicated in greyed boxes. Altogether, out of the 405 collected samples (from 142 SARS-CoV-2-positive patients), 232 samples from 130 patients with a documented date for the SARS-CoV-2-specific PCR-positive test, and 105 samples from 63 patients with a documented date of symptom onset were tested with both VIDAS^®^ SARS-CoV-2 IgM and IgG assays (paired tests).

For the evaluation of serum cross-reactivity, up to 276 frozen pre-pandemic sera (i.e. negative for SARS-CoV-2) collected from patients with other potentially interfering infections or medical conditions (bioMérieux, Centre Hospitalier Universitaire Grenoble-Alpes and St Joseph St Luc Lyon collections) were tested with the VIDAS^®^ SARS-CoV-2 IgM (276 sera from 33 medical conditions; one to 30 sera per condition) and the VIDAS^®^ SARS-CoV-2 IgG (261 sera from 33 medical conditions; two to 30 sera per condition) assays.

### Serological assays

VIDAS^®^ SARS-CoV-2 IgM (423833) and VIDAS^®^ SARS-CoV-2 IgG (423834) (bioMérieux, France) are automated qualitative CE-IVD assays developed for the VIDAS^®^ family of instruments and based on a two-step enzyme immunoassay combined with an enzyme-linked fluorescent assay (ELFA) detection technique. The VIDAS^®^ SARS-CoV-2 IgM and VIDAS^®^ SARS-CoV-2 IgG assays are intended for use as an aid to determine if individuals may have been exposed and infected by SARS-CoV-2 and if they have mounted a specific anti-SARS-CoV-2 IgM and IgG immune response. These assays allow the detection of SARS-CoV-2-specific IgM and IgG, respectively, from 100 μl serum or plasma (lithium heparin). The VIDAS^®^ SARS-CoV-2 IgM and VIDAS^®^ SARS-CoV-2 IgG serological assays were conducted according to the manufacturer’s instructions.

Briefly, a solid-phase receptacle coated with the antigen (recombinant SARS-CoV-2 receptor-binding domain [RBD] of the viral Spike protein) serves as both solid phase and pipetting device. After the sample dilution step, SARS-CoV-2-specific IgM and IgG are captured on the coated antigen and unbound components are washed out. In a second step, human IgM (VIDAS^®^ SARS-CoV-2 IgM) or IgG (VIDAS^®^ SARS-CoV-2 IgG) are specifically detected by mouse monoclonal antibodies conjugated to alkaline phosphatase and directed against human IgM or IgG, respectively. Unbound components are eliminated by washing and detection is performed by incubation with the substrate (4-Methyl-umbelliferyl phosphate) followed by measurement of the fluorescent product (4-Methyl-umbelliferone) at 450 nm. A relative fluorescence value (RFV) is generated (background reading subtracted from the final fluorescence reading). The assay is conducted with a standard (S1) and a positive control (C1) that contains humanized recombinant anti-SARS-CoV-2 antibody, either IgM or IgG depending on the assay. A negative control (C2) is also supplied. The results are automatically calculated by the instrument, according to the S1 standard, and an index value (i) is obtained (where i=RFV_sample_/RFV_S1_). The test is interpreted as negative when i < 1.00 and positive when i ≥ 1.00. The positivity cut-off values for the IgM and IgG tests were determined from a healthy pre-pandemic cohort (259 [IgM test] and 120 [IgG test] samples collected prior to August 2019), using non-parametric 99^th^ percentile because of normality rejection for the IgM positivity cut-off and using the (99,99) tolerance intervals approach after Box-Cox transformation (24) for the IgG positivity cut-off (99^th^ percentile index values at a 99% confidence level) (data not shown).

### Statistical analysis

Assay precision was evaluated according to the Clinical & Laboratory Standards Institute (CLSI) EP05-A3 guideline (25) by the variance component method and restricted maximum likelihood (REML) using the SAS Enterprise Guide 7.13 HF8 software.

Specificity and sensitivity (PPA) estimates were evaluated according to the CLSI EP12-A2 guideline (26). The 95% confidence intervals (95% CI) were computed (either as score confidence interval if the specificity or sensitivity (PPA) belonged to] 5; 95 [%, or as exact confidence interval otherwise) using the SAS Enterprise Guide 7.13 HF8 software.

PPA was evaluated per time windows (in days) relative to the day of RT-PCR positive result and of symptom onset (when documented). To avoid a statistical bias, only one patient’s measurement per time period was included in the analysis. In case of multiple patient’s measurements in one period, the first available measurement was considered for the calculation. Therefore, depending on the total number of longitudinal tests performed, each patient contributed with one to five test results in the five time windows considered (0-7, 8-15, 16-23, 24-31, ≥ 32 days).

The positive predictive value (PPV) and the negative predictive value (NPV) were calculated assuming a prevalence of 5%, as recommended by FDA for the EUA application (22), and the respective 95% CI were computed according to Mercaldo et al. (27) using the SAS Enterprise Guide 7.13 HF8 software.

VIDAS^®^ SARS-CoV-2 index values were displayed per time frame as Tukey box plots. Two-group comparisons of index values per time frame between hospitalized and non-hospitalized patients were performed using the nonparametric two-tailed Mann-Whitney *U*-test (MWU-test) with normal approximation. In case of multiple group comparisons, the Bonferroni method was applied for controlling the 5% overall probability of a false-significant result. Accordingly, for the three-group comparison shown in Figure 3, *P*-values < 0.017 were considered statistically significant.

## RESULTS

### Analytical performance of VIDAS ^®^ SARS-CoV-2 IgM and IgG assays

Within-run and within-laboratory precisions of the ELFA-based tests were determined using three samples (one negative and two positive for SARS-CoV-2 IgM and IgG). Samples were run in triplicate on one VIDAS^®^ instrument, twice a day over 10 days (with an instrument calibration every second day), using one assay lot, thus generating 60 measurement values per sample. The coefficient of variation (%CV) for repeatability (within-run precision) did not exceed 9.3% and 5.9% for the VIDAS^®^ SARS-CoV-2 IgM and IgG assays, respectively. The %CV for within-laboratory precision was also low, reaching a maximum of 10.7% and 6.9% for the VIDAS^®^ SARS-CoV-2 IgM and IgG assays, respectively (Table S1).

Analytical specificity and sensitivity of the VIDAS^®^ SARS-CoV-2 IgM and IgG assays were verified through various experiments. First, we ruled out a possible cross-reactivity of the anti-human-IgM (VIDAS^®^ SARS-CoV-2 IgM) or of the anti-human-IgG (VIDAS^®^ SARS-CoV-2 IgG) with human IgG or IgM, respectively, which might produce false-positive results. Spike-in experiments in negative samples using either human recombinant monoclonal anti-SARS-CoV-2 IgG (10 μg/ml) in the VIDAS^®^ SARS-CoV-2 IgM assay or human recombinant monoclonal anti-SARS-CoV-2 IgM (3 μg/ml) in the VIDAS^®^ SARS-CoV-2 IgG assay demonstrated neither reactivity of the alkaline-phosphatase-conjugated anti-human-IgM toward human IgG nor reactivity of the anti-human-IgG toward human IgM (n=10; data not shown). Second, we ruled out a possible competition between anti-SARS-CoV-2 IgM and IgG for binding to the coated SARS-CoV-2 antigen, which might interfere with the respective assays and generate false-negative results. Spike-in experiments in positive samples (with index values ranging from 1.8 to 20.6) using an excess of human recombinant monoclonal anti-SARS-CoV-2 IgG (10 μg/ml) in the VIDAS^®^ SARS-CoV-2 IgM assay or of human recombinant monoclonal anti-SARS-CoV-2 IgM (3 μg/ml) in the VIDAS^®^ SARS-CoV-2 IgG assay did not impact the qualitative test results and resulted in a maximum deviation from the respective control of 17.6% (n=12; data not shown). Third, we evaluated the impact of serum inactivation (56°C for 30 minutes), which might be applied by diagnostics laboratories to inactivate potentially infectious samples (28), on test results of 10 negative and 10 positive samples. Heat inactivation did not impact the qualitative test results of the VIDAS^®^ SARS-CoV-2 IgM and IgG assays (20/20 = 100% concordance) and the maximum deviation to the (non-heated) control among 18/20 samples was 17%. However, one negative sample (VIDAS^®^ SARS-CoV-2 IgG) and one positive sample (VIDAS^®^ SARS-CoV-2 IgM) yielded significantly divergent index values (data not shown). Therefore heat inactivation of sera prior to VIDAS^®^ SARS-CoV-2 testing should be preferably avoided. Finally, we evaluated the possible cross-reactivity of components of the assay (SARS-CoV-2 antigen RBD or immunoglobulins) with human sera from patients with other infections (including other coronaviruses) or medical conditions (e.g. rheumatoid factor) (29) that might interfere with the assay and yield false-positive results. Up to 276 (VIDAS^®^ SARS-CoV-2 IgM) and 261 (VIDAS^®^ SARS-CoV-2 IgG) sera of SARS-CoV-2-negative patients with other infections or conditions were tested and the number of positive test results was evaluated (Table 1). None of the 18 sera of patients with an history of infection with the human coronaviruses CoV-NL63, CoV-229E, CoV-HKU1 or CoV-OC43 (genera *Alphacoronavirus* and *Betacoronavirus*) were positive in the VIDAS^®^ SARS-CoV-2 IgG assay, while the serum of one CoV-NL63-positive patient was positive in the VIDAS^®^ SARS-CoV-2 IgM assay. Only two out of 261 (0.8%) tested sera were positive in the VIDAS^®^ SARS-CoV-2 IgG assay. They belonged to a HIV-positive and a respiratory syncytial virus (RSV A)-positive patient, respectively. On the other hand, ten out of 276 (3.6%) tested sera were positive in the VIDAS^®^ SARS-CoV-2 IgM assay. Apart from the one CoV-NL63-positive sample mentioned above, six sera were from patients presenting autoantibodies (antinuclear antibody, rheumatoid factor), two were from patients with an history of parasite infection (*Plasmodium falciparum, Trypanosoma cruzi*), and one from a past Rhinovirus/Enterovirus infection. The index values associated with these 12 crossreactive sera was low, with a median (interquartile range) of 2.1 (1.4-3.8). None of the sera from patients infected with other respiratory viruses, including influenza virus, parainfluenza virus, metapneumovirus or adenovirus were reactive.

**Table 1.** Cross-reactivity of human sera from patients with other infections or medical conditions potentially interfering with the VIDAS ^®^ SARS-CoV-2 IgM and IgG assays

| Sample category | VIDAS® SARS-CoV-2 IgM |  | VIDAS® SARS-CoV-2 IgG |  |
| --- | --- | --- | --- | --- |
|  | No. samples tested | No. positive tests | No. samples tested | No. positive tests |
| Pregnant women | 5 | 0 | 5 | 0 |
| Antinuclear antibody (ANA)<sup>a</sup> | 47 | 2 | 47 | 0 |
| Rheumatoid factor | 19 | 4 | 19 | 0 |
| Human anti-mouse antibody (HAMA) | 5 | 0 | 5 | 0 |
| *Borrelia burgdorferi*<sup>b</sup> | 10 | 0 | 6 | 0 |
| *Haemophilus Influenzae B* | 5 | 0 | 5 | 0 |
| *Plasmodium falciparum* | 3 | 1 | 3 | 0 |
| *Toxoplasma gondii*<sup>b</sup> | 10 | 0 | 6 | 0 |
| *Treponema pallidum* | 3 | 0 | 3 | 0 |
| *Trypanosoma cruzi* | 5 | 1 | 5 | 0 |
| Hepatitis A virus (HAV) | 3 | 0 | 3 | 0 |
| Hepatitis B virus (HBV) | 5 | 0 | 5 | 0 |
| Hepatitis C virus (HCV) | 5 | 0 | 5 | 0 |
| Hepatitis E virus (HEV)<sup>b</sup> | 7 | 0 | 6 | 0 |
| Herpes simplex virus (HSV)<sup>b</sup> | 6 | 0 | 6 | 0 |
| Human immunodeficiency virus (HIV) | 5 | 0 | 5 | 1 |
| Cytomegalovirus (CMV) | 4 | 0 | 3 | 0 |
| Measles virus (MV) | 4 | 0 | 3 | 0 |
| Mumps virus (MuV) | 1 | 0 | 3 | 0 |
| Rubella virus (RuV)<sup>b</sup> | 10 | 0 | 6 | 0 |
| Dengue virus (DENV) | 3 | 0 | 3 | 0 |
| West Nile virus (WNV) | 4 | 0 | 3 | 0 |
| Yellow fever virus (YFV) | 4 | 0 | 3 | 0 |
| Zika virus (ZIKV)<sup>b</sup> | 5 | 0 | 5 | 0 |
| Adenovirus (AdV) | 2 | 0 | 2 | 0 |
| Metapneumovirus (MPV) | 4 | 0 | 4 | 0 |
| Rhinovirus/Enterovirus (RV/Enterov)<sup>c</sup> | 20 | 1 | 20 | 0 |
| Influenza A and B virus (IAV/IBV) | 30 | 0 | 30 | 0 |
| Parainfluenza viruses 1/2/3 (PIV-1/2/3) | 11 | 0 | 11 | 0 |
| Respiratory syncytial virus A or B (RSV A or B) | 13 | 0 | 13 | 1 |
| Coronavirus NL63/HKU1 (CoV-NL63/HKU1)<sup>d</sup> | 9 | 1 | 9 | 0 |
| Coronavirus 229E (CoV-229E) | 7 | 0 | 7 | 0 |
| Coronavirus OC43 (CoV-OC43) | 2 | 0 | 2 | 0 |
| **Total** | **276** | **10** | **261** | **2** |

<sup>a</sup>Include anti-DNA, anti-SSA, anti-SSB and anti-Sm/RNP antibodies; <sup>b</sup>The proportion of acute infection (i.e. IgM-positive for the respective infectious agent, when IgM levels were characterized) among samples tested with the VIDAS® SARS-CoV-2 IgM assay was: *Borrelia burgdorferi*: 5/10, *Toxoplasma gondii*: 5/10, HEV: 3/7, HSV: 3/6, RuV: 5/10, ZIKV: 3/5; <sup>c</sup>One out of 20 sera was from a patient with a bocavirus (BoV) co-infection and was negative in both VIDAS® SARS-CoV-2 IgM and IgG assays; <sup>d</sup>Six out of the nine sera tested were from patients positive for CoV-NL63 and three were from patients positive for CoV-NL63 and/or CoV-HKU1.

### Clinical performance of VIDAS ^®^ SARS-COV-2 IgM and IgG assays

The clinical specificity of the VIDAS^®^ SARS-CoV-2 IgM and IgG assays were assessed using sera from up to 989 pre-pandemic healthy volunteers collected in France and in the United States before September 2019. A total of 308 sera were tested with the VIDAS^®^ SARS-CoV-2 IgM assay and 989 sera were tested with the VIDAS^®^ SARS-CoV-2 IgG assay. The combined IgM/IgG assay specificity (defined as both VIDAS^®^ SARS-CoV-2 IgM and IgG assays being negative) was determined on 308 paired VIDAS^®^ SARS-CoV-2 IgM and IgG tests (Tables S2 and S3). 306/308 (VIDAS^®^ SARS-CoV-2 IgM) and 988/989 (VIDAS^®^ SARS-CoV-2 IgG) SARS-CoV-2-negative sera were negative, corresponding to a specificity (95% CI) of 99.4% (97.7-99.9%) and 99.9% (99.4-100%) for the VIDAS^®^ SARS-CoV-2 IgM and IgG assays, respectively (Table S3). The specificity (95% CI) of the VIDAS^®^ SARS-CoV-2 IgG test on the common cohort (N=308) was 100.0% (98.8-100.0%). The specificity (95% CI) of the combined IgM and IgG serology tests (306/308 tests negative in both assays) was 99.4% (97.7-99.9%) (Table S3).

The clinical sensitivity of the VIDAS^®^ SARS-CoV-2 IgM and IgG assays was assessed using 405 samples collected from 142 patients confirmed positive for SARS-CoV-2 by RT-PCR. The positive percent agreement (PPA) of the serology tests with the RT-PCR test results was calculated per weekly time windows (0-7, 8-15, 16-23, 24-31 and ≥ 32 days) relative to the time from the PCR-positive result and to the time from symptom onset. No more than one patient sample per time window was included in the calculation (Fig. 1 and Table 2). The PPA calculated on all available samples for the VIDAS^®^ SARS-CoV-2 IgM and IgG assays are shown in Tables S4 and S5, respectively. For the sake of comparability, the PPA (95% CI) of the IgM, IgG and combined IgM/IgG test results was also calculated on samples evaluated with both tests (paired testing; Tables 3, 4 and S6).

**Table 2.** Demographics of French patients positive for SARS-CoV-2 used for the determination of VIDAS ^®^ SARS-CoV-2 IgM and IgG sensitivity (positive percent agreement with RT-PCR positivity)

| VIDAS® SARS-CoV-2 serology testing | IgM | IgG | Combined IgM/IgG |
| --- | --- | --- | --- |
| **Patients with documented date of RT-PCR-positive test** |  |  |  |
| Number of samples tested | 234 | 253 | 232 |
| Study population, N (%) | 132 (100%) | 139 (100%) | 130 (100%) |
| Age in years, median (range) |  |  |  |
| Non-hospitalized (N=61) | missing | missing | missing |
| Other (N=71/78/69) | 71 (27-96) | 70.5 (27-96) | 70 (27-96) |
| Gender, N (%) |  |  |  |
| Male | 47 (35.6%) | 53 (38.1%) | 47 (36.2%) |
| Female | 24 (18.2%) | 25 (18.0%) | 22 (16.9%) |
| Missing | 61 (46.2%) | 61 (43.9%) | 61 (46.9%) |
| Disease severity, N (%) |  |  |  |
| Hospitalized | 56 (42.4%) | 57 (41.0%) | 54 (41.55%) |
| Non-hospitalized | 61 (46.2%) | 61 (43.9%) | 61 (46.9%) |
| Missing | 15 (11.4%) | 21 (15.1%) | 15 (11.55%) |
| **Patients with documented date of symptom onset** |  |  |  |
| Number of samples tested | 108 | 113 | 105 |
| Study population, N (%) | 65 (100%) | 71 (100%) | 63 (100%) |
| Age in years, median (range) | 70 (27-96) | 70 (27-96) | 70 (27-96) |
| Gender, N (%) |  |  |  |
| Male | 45 (69.2%) | 51 (71.8%) | 45 (71.4%) |
| Female | 20 (30.8%) | 20 (28.2%) | 18 (28.6%) |
| Disease severity, N (%) |  |  |  |
| Hospitalized | 50 (76.9%) | 50 (70.4%) | 48 (76.2%) |
| Non-hospitalized<sup>a</sup> | 0 (0.00%) | 0 (0.0%) | 0 (0.0%) |
| Missing | 15 (23.1%) | 21 (29.6%) | 15 (23.8%) |

<sup>a</sup>Date of symptom onset not documented in non-hospitalized COVID-19 patients

**Table 3.** Positive percent agreement (PPA) of the VIDAS ^®^ SARS-CoV-2 IgM, IgG and combined IgM/IgG test results of SARS-CoV-2-positive samples, according to the time from RT-PCR-positive result

| VIDAS<sup>®</sup> SARS-CoV-2 serology testing | Time from RT-PCR-positive result | Median (range) time in days | No. samples<sup>a</sup> | No. positive results | PPA | 95% CI |
| --- | --- | --- | --- | --- | --- | --- |
| **IgM (N=232)** | 0-7 days | 2 (0-7) | 110 | 49 | 44.5% | 35.6-53.9% |
|  | 8-15 days | 14 (8-15) | 60 | 49 | 81.7% | 70.1-89.4% |
|  | 16-23 days | 20 (16-23) | 38 | 31 | 81.6% | 66.6-90.8% |
|  | 24-31 days | 26 (24-28) | 13 | 13 | 100.0% | 75.3-100.0% |
|  | ≥ 32 days | 33 (32-65) | 11 | 9 | 81.8% | 52.3-94.9% |
| **IgG (N=232)** | 0-7 days | 2 (0-7) | 110 | 50 | 45.5% | 36.5-54.8% |
|  | 8-15 days | 14 (8-15) | 60 | 53 | 88.3% | 77.8-94.2% |
|  | 16-23 days | 20 (16-23) | 38 | 36 | 94.7% | 82.7-98.5% |
|  | 24-31 days | 26 (24-28) | 13 | 13 | 100.0% | 75.3-100.0% |
|  | ≥ 32 days | 33 (32-65) | 11 | 11 | 100.0% | 71.5-100.0% |
| **Combined IgM/IgG<sup>b</sup> (N=232)** | 0-7 days | 2 (0-7) | 110 | 59 | 53.6% | 44.4-62.7% |
|  | 8-15 days | 14 (8-15) | 60 | 57 | 95.0% | 86.1-99.0% |
|  | 16-23 days | 20 (16-23) | 38 | 38 | 100.0% | 90.7-100.0% |
|  | 24-31 days | 26 (24-28) | 13 | 13 | 100.0% | 75.3-100.0% |
|  | ≥ 32 days | 33 (32-65) | 11 | 11 | 100.0% | 71.5-100.0% |

<sup>a</sup>no more than one test result per patient per time period; <sup>b</sup>combined test is positive when at least one of IgM and/or IgG tests are positive

**Table 4.**
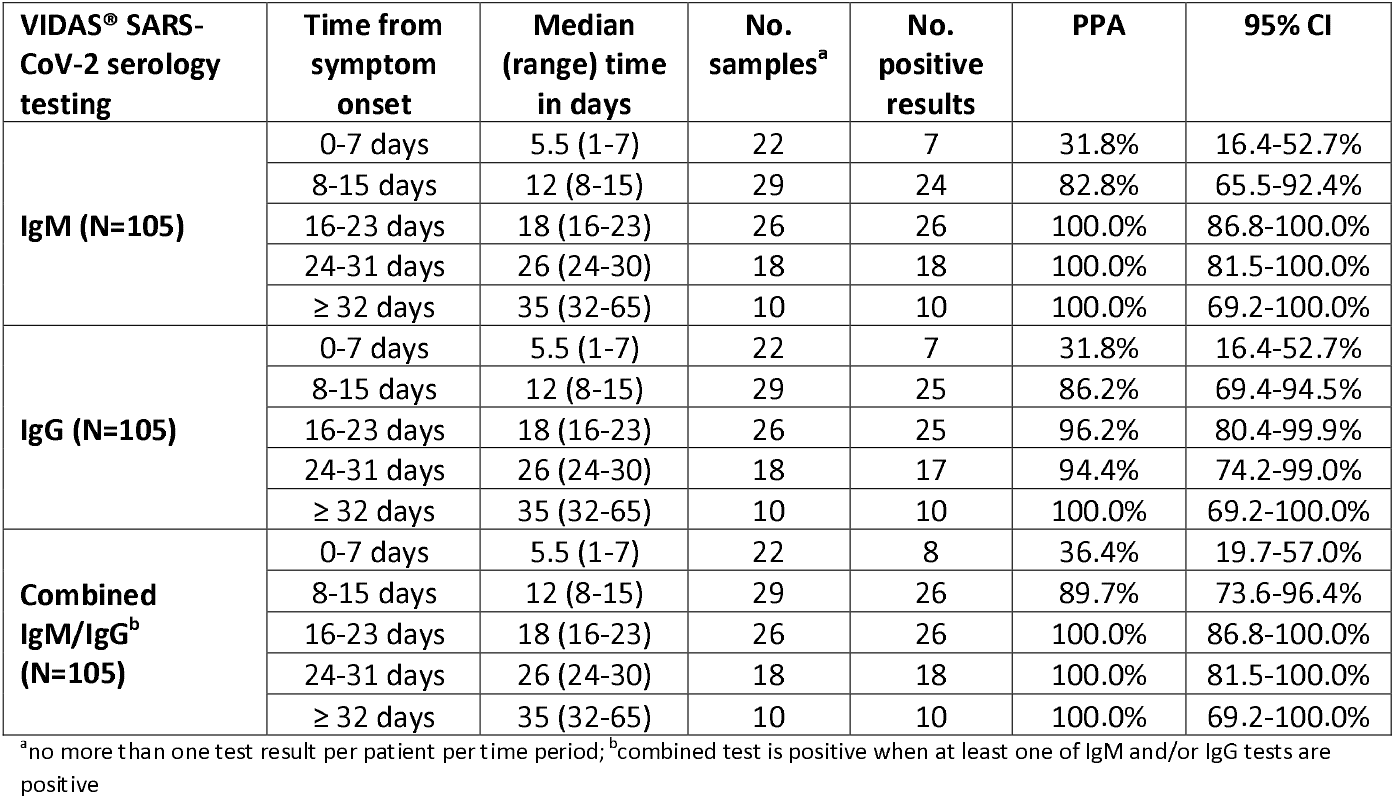
Positive percent agreement (PPA) of the VIDAS ^®^ SARS-CoV-2 IgM, IgG and combined IgM/IgG test results of SARS-CoV-2-positive samples, according to the time from symptom onset

The PPA from the time of RT-PCR-positive test results for the VIDAS^®^ SARS-CoV-2 IgM assay raised from 44.5% at 0-7 days to 100.0% at 24-31 days, before decreasing to 81.8% at ≥ 32 days (Table 3). The PPA for the VIDAS^®^ SARS-CoV-2 IgG assay raised from 45.5% at 0-7 days to 100.0% from day 24 onwards(Table 3). The PPA of the combined IgM and IgG serology tests (positive in at least one of the IgM and/or IgG assays) from the time of RT-PCR-positive test results, increased from 53.6% at 0-7 days to 100.0% from day 16 onwards (Tables 3 and S6). The PPA evaluated relative to the time of symptom onset raised from 31.8% at 0-7 days (VIDAS^®^ SARS-CoV-2 IgM and IgG) to 100.0% from day 16 (VIDAS^®^ SARS-CoV-2 IgM) or day 24 (VIDAS^®^ SARS-CoV-2 IgG) (Table 4). The PPA of the combined IgM and IgG serology test results from the time of symptom onset, increased from 36.4% at 0-7 days to 100.0% from day 16 onwards (Tables 4 and S6). It should be noted that the PPA relative to the date of symptom onset is mainly based on hospitalized patients (Table 2).

Based on the specificity and the PPA determined on paired IgM and IgG testing (N=308 for specificity; N=105 for PPA), the negative predictive value (NPV) and the positive predictive value (PPV) were calculated at 5% prevalence (22), according to the time after symptom onset (Table 5). The NPV was high for both the VIDAS^®^ SARS-CoV-2 IgM and IgG assays, whether considered alone or in combination; NPV was ≥ 96.5% (lower 95% confidence limits ≥ 95.4%) at 0-7 days post symptom onset and NPV increased from 99.1% to 100.0% (lower 95% confidence limits ≥ 98.0%) from day 8 onwards following symptom onset. The PPV of the VIDAS^®^ SARS-CoV-2 IgG assay was 100% at all time frames considered. Of note, the PPV calculated using the full data set for the VIDAS^®^ SARS-CoV-2 IgG specificity determination (N=989) was slightly lower, increasing from 94.3% at 0-7 days to 98.1% at ≥ 32 days (Table S7). The PPV of the VIDAS^®^ SARS-CoV-2 IgM assay was lower, ranging from 72.1% at 0-7 days to 89.0% from day 16 onwards following symptom onset (Table 5). The combination of IgM and IgG test results slightly improved the PPV and NPV of the SARS-CoV-2 IgM assay, and the NPV of the SARS-CoV-2 IgG assay. The SARS-CoV-2 IgG assay performed best alone in terms of PPV (Table 5).

**Table 5.** Positive and negative predictive values (PPV/NPV) at 5% prevalence of the VIDAS ^®^ SARS-CoV-2 IgM, IgG and combined IgM/IgG test results, according to the time from symptom onset

| VIDAS® SARS-CoV-2 serology testing | Time from symptom onset | PPV<sup>a</sup> (95% CI) | NPV<sup>a</sup> (95% CI) |
| --- | --- | --- | --- |
| **IgM** | 0-7 days | 72.1% (36.3-92.1%) | 96.5% (95.4-97.4%) |
|  | 8-15 days | 87.0% (62.5-96.4%) | 99.1% (98.0-99.6%) |
|  | 16-23 days | 89.0% (67.1-97.0%) | 100.0% (N/A)<sup>b</sup> |
|  | 24-31 days | 89.0% (67.1-97.0%) | 100.0% (N/A)<sup>b</sup> |
|  | ≥ 32 days | 89.0% (67.1-97.0%) | 100.0% (N/A)<sup>b</sup> |
| **IgG** | 0-7 days | 100.0% (N/A)<sup>b</sup> | 96.5% (95.4-97.4%) |
|  | 8-15 days | 100.0% (N/A)<sup>b</sup> | 99.3% (98.2-99.7%) |
|  | 16-23 days | 100.0% (N/A)<sup>b</sup> | 99.8% (98.6-100.0%) |
|  | 24-31 days | 100.0% (N/A)<sup>b</sup> | 99.7% (98.1-100.0%) |
|  | ≥ 32 days | 100.0% (N/A)<sup>b</sup> | 100.0% (N/A)<sup>b</sup> |
| **Combined IgM/IgG** | 0-7 days | 74.7% (40-92.9%) | 96.7% (95.6-97.6%) |
|  | 8-15 days | 87.9% (64.5-96.7%) | 99.5% (98.4-99.8%) |
|  | 16-23 days | 89.0% (67.1-97.0%) | 100.0% (N/A)<sup>b</sup> |
|  | 24-31 days | 89.0% (67.1-97.0%) | 100.0% (N/A)<sup>b</sup> |
|  | ≥ 32 days | 89.0% (67.1-97.0%) | 100.0% (N/A)<sup>b</sup> |

<sup>a</sup>PPV and NPV were calculated at 5% prevalence and using values of specificity and sensitivity (PPA) determined on paired VIDAS® SARS-CoV-2 IgM and IgG test results (N=308 for specificity, Table S3; N=105 for PPA, Table 4); <sup>b</sup>95% CI not calculable (division by zero)

### Longitudinal study of IgM and IgG seroconversion in hospitalized COVID-19 patients

The global distribution of VIDAS^®^ SARS-CoV-2 IgM and IgG index values post symptom onset was compared among the 105 SARS-CoV-2-positive patients described in Table 4 (i.e. mainly including hospitalized COVID-19 patients; Table 2) (Fig. 2). IgM index values increased from the second week of symptom onset (8-15 days) and peaked during the third week (16-23 days) before decreasing. In comparison, the IgG index values strongly increased from the second week of symptom onset and seemed to reach a plateau ≥ 32 days post symptom onset (Fig. 2).

**Figure 2.**
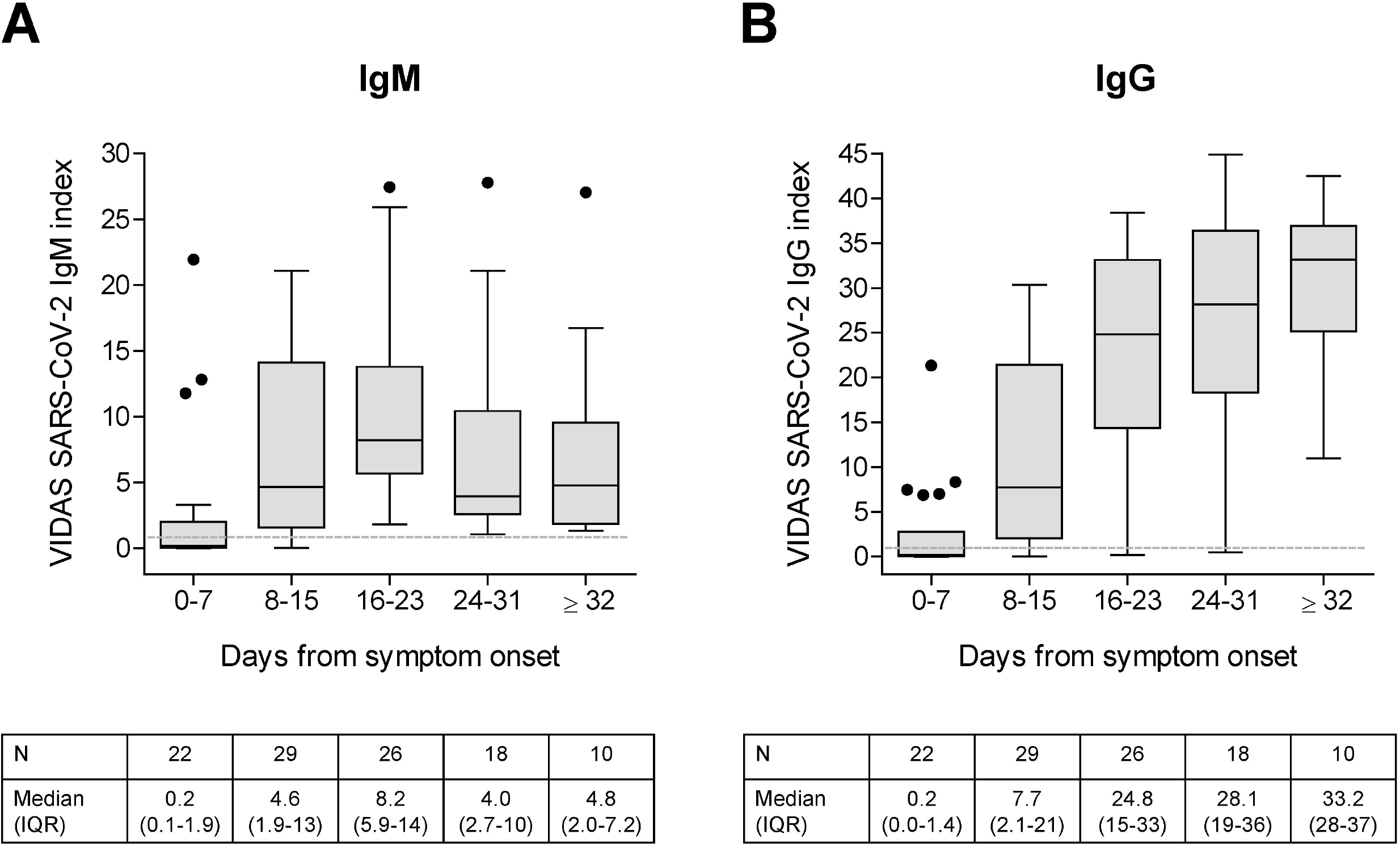
Distribution of IgM (A) and IgG (B) index values obtained using the VIDAS ^®^ SARS-CoV-2 IgM and IgG assays, respectively, in patients confirmed positive for SARS-CoV-2, according to the time from symptom onset. VIDAS^®^ SARS-CoV-2 IgM and IgG index values of 105 paired tests from 63 SARS-CoV-2-positive patients (Table 4, Table 2 and Fig. 1) are displayed as Tukey box plots according to the time from symptom onset. No more than one patient’s sample is included per time frame. The number of tested samples (N), and the median and interquartile range (IQR) of index values are indicated below each graph. The dashed line shows the positivity cut-off of both assays (i = 1.00).

The VIDAS^®^ SARS-CoV-2 IgM and IgG index values were also compared between hospitalized and non-hospitalized patients. Since the date of symptom onset was not documented in non-hospitalized patients (Table 2), the index values in 54 hospitalized and 61 non-hospitalized patients were compared relative to the time of the RT-PCR-positive results (Fig. 3). Interestingly, the distribution of index values between hospitalized and non-hospitalized patients differ statistically from each other for both the VIDAS^®^ SARS-CoV-2 IgM and IgG assays at the three compared time frames (0-7, 8-15 and 16-23 days post PCR-positive test; MWU-test p-values < 0.017). Median index values were higher in hospitalized versus non-hospitalized patients (Fig. 3).

**Figure 3.**
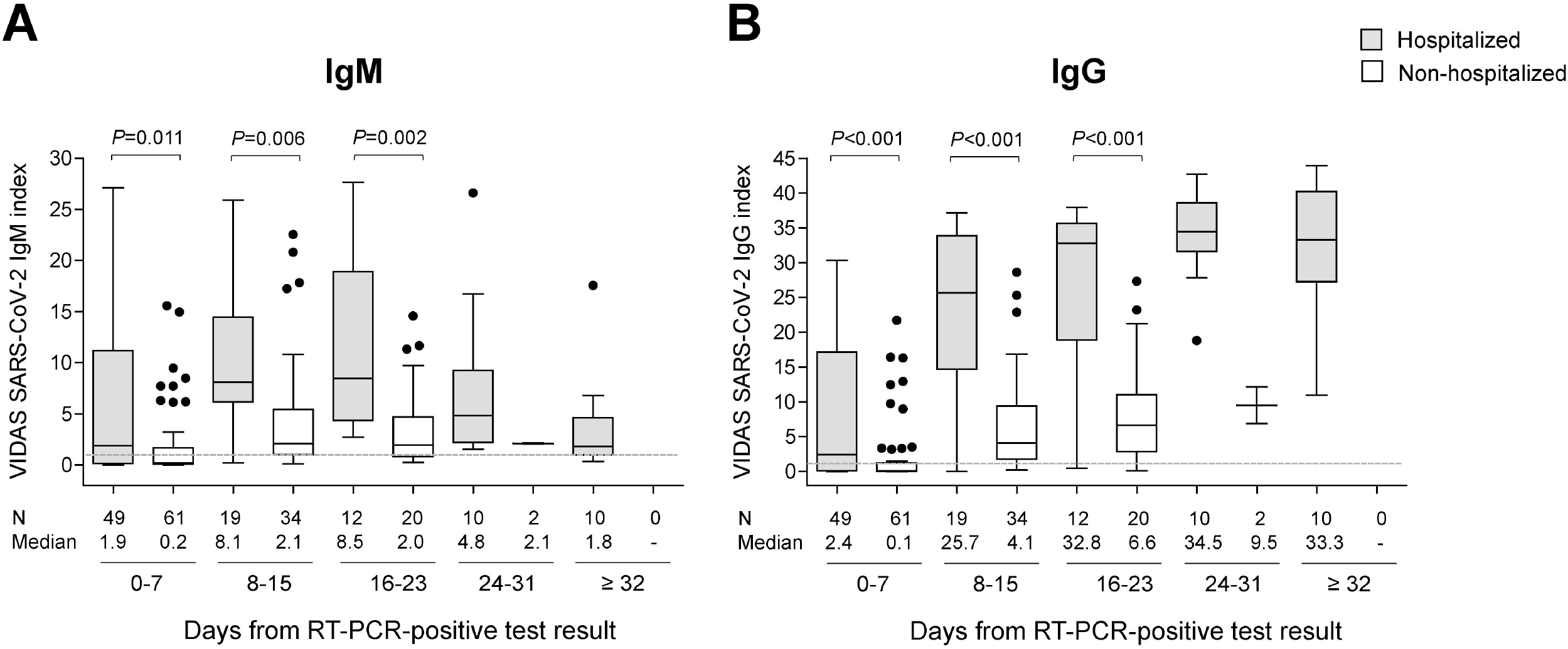
Distribution of IgM (A) and IgG (B) index values obtained using the VIDAS ^®^ SARS-CoV-2 IgM and IgG assays, respectively, in hospitalized vs. non-hospitalized patients confirmed positive for SARS-CoV-2, according to the time from PCR-positive result. Out of the 232 paired tests (Table 3), 15 were from patients with an unknown hospitalization status (Table 2) and were thus excluded from the analysis. VIDAS^®^ SARS-CoV-2 IgM and IgG index values of 217 paired tests from 115 SARS-CoV-2-positive patients (100 samples from 54 hospitalized patients and 117 samples from 61 non-hospitalized patients) are depicted as Tukey box plots according to the time from RT-PCR-positive test result. No more than one patient’s sample is included per time frame. The number of tested samples (N) and the median index are indicated below each graph. The dashed line shows the positivity cut-off of both assays (i = 1.00). Differences between the groups of hospitalized and non-hospitalized patients were tested at each time frame (0-7, 8-15, 16-23 days post positive PCR) using a two-sided MWU-test; the respective p-values are displayed above each graph. No statistical testing was performed at 24-31 and ≥ 32 days due to the too low N values, notably in the group of non-hospitalized patients.

Finally, IgM and IgG seroconversion was further investigated in four selected hospitalized patients with either early and/or repeated measurements over an extended period of time (up to 74 days post symptom onset; Fig. 4). IgG seroconversion closely followed IgM seroconversion in the second week of symptom onset (Fig. 4A-B), in line with the global profile shown in Figure 2. SARS-CoV-2 IgM index rapidly decreased concomitantly with the increase of SARS-CoV-2 IgG index (Fig. 4B-D). In the three patients shown in Fig. 4B-D, SARS-CoV-2 IgM index decreased below the positivity cutoff (index = 1.00) 46 days after symptom onset. In contrast, SARS-CoV-2 IgG index values remained high and stable from approximately day 20 onward after symptom onset, at least up to 74 days.

**Figure 4.**
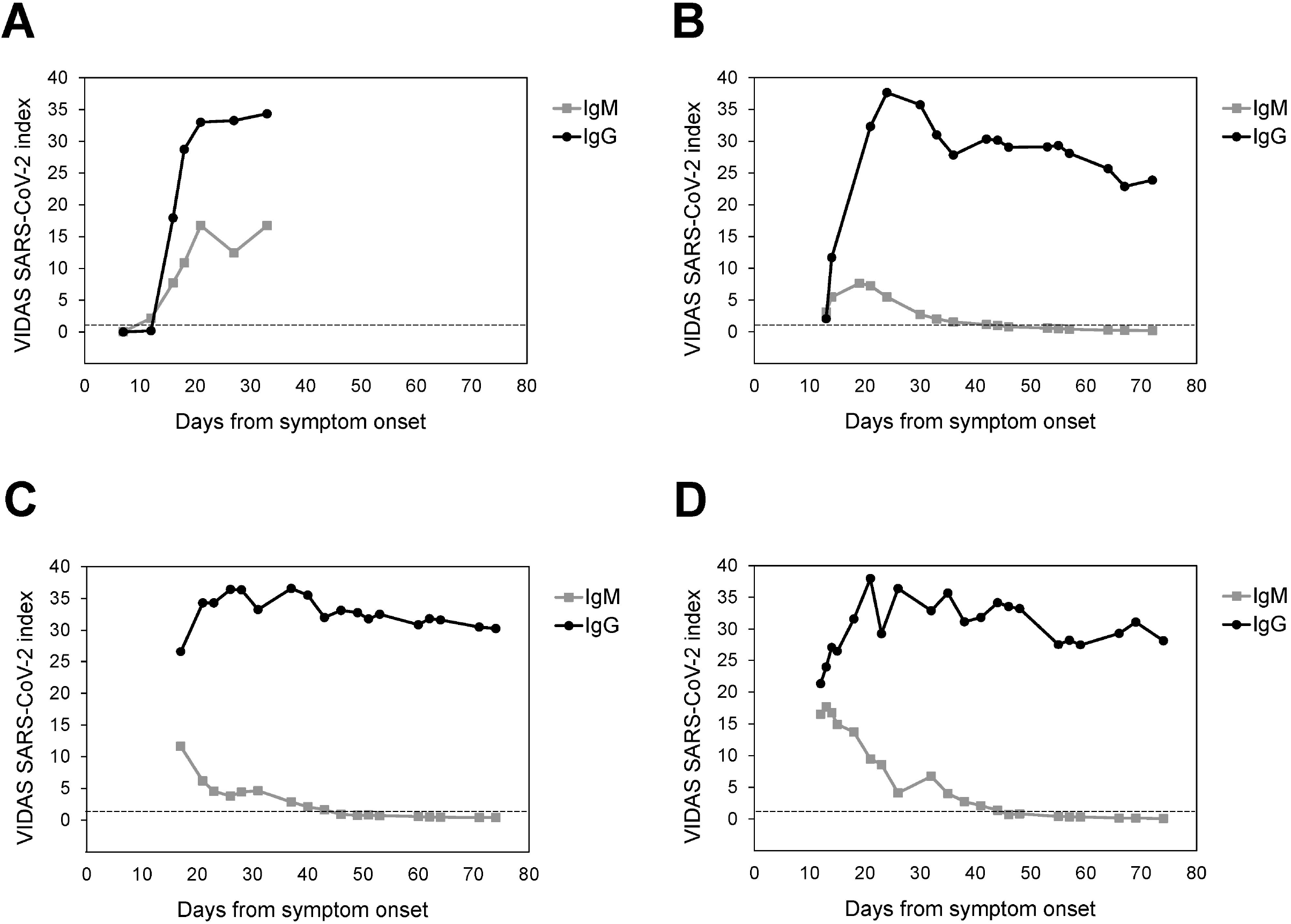
Kinetics of IgM and IgG seroconversion in four selected hospitalized patients. VIDAS^®^ SARS-CoV-2 IgM and IgG index values of four patients (A to D, respectively) measured over time after symptom onset are presented. The dashed line indicates the positivity cut-off of both assays (i = 1.00). Further patients’ information is as follows: (**A**) the 78-year-old male patient was in the intensive-care unit (ICU) at all investigated time points, except at the first (day 7) and last (day 33) measurement time points; (**B**) the 77-year-old male patient was in ICU at all investigated time points; (**C**) the 43-year-old male patient was in ICU at all investigated times, except at the last two measurement time points (day 71 and 74); (**D**) the 67-year-old male patient was in ICU at all investigated time points.

## DISCUSSION

We describe here the analytical and clinical performance of the VIDAS^®^ SARS-CoV-2 IgM and IgG assays. We demonstrate that both assays show high precision, and excellent analytical and clinical performances.

The rate of cross-reactivity with non-specific antibodies, including those of patients infected with other coronaviruses, was very low in both VIDAS^®^ SARS-CoV-2 IgM and IgG assays. This weak cross-reactivity with other coronaviruses antibodies is likely due, at least in part, to the choice of the receptor-binding domain (RBD) of the viral Spike protein as SARS-CoV-2-specific antigen. The RBD shows a high sensitivity in ELISA, higher than that of the SARS-CoV-2 spike S1 or nucleocapsid (NC) antigens (30–33). It also presents a weaker homology and significant structural divergences with the RBD of other coronaviruses (34–36). Another advantage of using the RBD is that the viral antigen generates neutralizing antibodies likely to provide protective immunity (35–42), as previously demonstrated for SARS-CoV (43, 44). That the VIDAS^®^ SARS-CoV-2 assays have the ability to detect SARS-CoV-2 neutralizing antibodies was recently demonstrated in mild COVID-19 patients, with an almost perfect concordance (Cohen’s Kappa coefficient of 0.9) between the VIDAS^®^ SARS-CoV-2 IgG assay and a virus neutralization test (42). Beside its strong immunogenicity and antigenicity, the RBD of SARS-CoV has been shown to elicit antibody responses that persisted many years after infection (38, 44), raising the possibility that it might also be the case for the RBD of SARS-CoV-2. Recent studies in COVID-19 patients, notably in convalescent donors (45–47), on the anti-RBD antibody dynamics post SARS-CoV-2 infection (39, 40), or demonstrating the persistence and expansion of SARS-CoV-2-specific memory lymphocytes (48), as well as the stability of the IgG response detected with the VIDAS^®^ SARS-CoV-2 IgG assay up to 74 days post-symptom onset in the present study, strongly support this proposition. Hence, a serology test such as the VIDAS^®^ SARS-CoV-2 IgG assay is likely to be suitable for the detection of protective immunity and the evaluation of the efficacy of future vaccines, which are mainly based on the RBD-containing Spike protein (49, 50).

The low cross-reactivity rate with non-specific sera probably explains the very high specificity (≥ 99%) and narrow 95% CI of both VIDAS^®^ SARS-CoV-2 IgM and IgG assays. The VIDAS^®^ SARS-CoV-2 IgG assay alone had a specificity close to 100%, slightly higher than that of the VIDAS^®^ SARS-CoV-2 IgM assay.

The clinical sensitivity of the VIDAS^®^ SARS-CoV-2 assays was evaluated in SARS-CoV-2-confirmed symptomatic cases and was determined as positive percent agreement (PPA) with the RT-PCR assay, at successive time frames post positive PCR and, alternatively, post symptom onset. The PPA reached 100% at 16-23 days (VIDAS^®^ SARS-CoV-2 IgM) and at ≥ 32 days (VIDAS^®^ SARS-CoV-2 IgG) post-symptom onset. The combined VIDAS^®^ SARS-CoV-2 IgM/IgG test evaluation improved the PPA of the respective IgM and IgG tests by 3.5 to 6.9 percent points during the first two weeks (0-7 and 8-15 days) of symptom onset. Such improved sensitivity of the combined IgM/IgG tests early after symptom onset might also be useful for the diagnosis of suspected COVID-19 cases with negative PCR (13–15, 17, 18).

Overall, the clinical performance of the VIDAS^®^ SARS-CoV-2 assays was excellent. It was in a range comparable to that reported for existing EUA serological assays (22, 33, 42, 51–57). Moreover, in several side-by-side comparisons of six to nine commercial serological assays (specific for SARS-CoV-2 IgA, IgM, IgG or total antibodies), VIDAS^®^ SARS-CoV-2 IgG outperformed some of the IgG-specific competitor assays in terms of specificity and/or PPA with PCR positivity (42, 56, 57). The high specificity of the VIDAS^®^ SARS-CoV-2 IgG assay alone should be well suited for epidemiological surveillance.

The kinetics of SARS-CoV-2 IgM and IgG seroconversion was also evaluated by monitoring VIDAS^®^ index values over time. VIDAS^®^ SARS-CoV-2 IgM and IgG index values increased in the second week after symptom onset. IgG index values strongly increased and remained high, as IgM index values rapidly declined. These profiles are in agreement with those described in recent publications (13, 30, 39, 40, 58–62). Interestingly, the magnitude of the antibody response (index values) correlated with disease severity, as it was significantly higher in hospitalized vs. non-hospitalized COVID-19 patients at the time frames investigated (0-7, 18-15 and 16-23 days after a PCR-positive test). This observation is in agreement with published reports (11, 40, 62–65). It should be noted that despite the significantly lower response of sera from mild (non-hospitalized) COVID-19 patients in the VIDAS^®^ SARS-CoV-2 assays (Fig. 3), the good performance of the assays as well as their strong concordance with seroneutralization was recently demonstrated in a cohort of mild COVID-19 patients (42).

This study presents several limitations. First, assay sensitivity was evaluated on confirmed but not on suspected SARS-CoV-2 cases (i.e. patients with symptoms but negative by PCR). It would be interesting to evaluate and confirm the benefit of SARS-CoV-2 IgM and IgG serology to complement PCR testing (13, 14, 17, 18). On the other hand, recent reports suggested that the identification rate of false-negative PCR results using serology testing might be marginal, between ∼1% (20) and ∼4% (62). Second, assay sensitivity was determined on symptomatic (hospitalized and non-hospitalized) COVID-19 patients. The sensitivity of the VIDAS^®^ SARS-CoV-2 IgM and IgG assays in asymptomatic SARS-CoV-2-infected individuals, who may represent most of the infected patients, remains to be evaluated.

In conclusion, VIDAS^®^ SARS-CoV-2 IgM and IgG are highly sensitive and specific assays for the reliable screening of patients after acute SARS-CoV-2 infections (and likely after vaccination, when available). Moreover, the VIDAS^®^ SARS-CoV-2 IgG assay fulfils the specificity requirement for its use in seroepidemiology studies and is well suited for the detection of past SARS-CoV-2 infections. Further studies are necessary to confirm its suitability for the detection of SARS-CoV-2 neutralizing antibodies and to define correlates of immune protection.

## Data Availability

All data are available upon request to bioMerieux.

## ACKNOWLEDGMENTS

This work was supported by bioMérieux. We thank the Etablissement Français du Sang, notably Yves Mérieux, for providing samples. We thank all the members of R&D Immunoassay bioMérieux who took part in this work, i.e. the bioMolecule Engineering team, Prototyping, Development and Verification teams. We warmly thank Nadia Piga and the biobank team for collecting the samples in a short time frame. We are grateful to the bioMérieux Data Sciences team for the calculations and statistical evaluation of the data, and to Victor Bondanese, Laurence Bridon and the Clinical Affairs team of bioMérieux for their help and technical support. We thank Karen Brengel for her useful advices as well as Alice Banz for critically reading the manuscript. The authors thank Dr. Anne Rascle of AR Medical Writing (Regensburg, Germany) for providing medical writing support, which was funded by bioMérieux (Marcy L’Etoile, France) in accordance with Good Publication Practice (GPP3) guidelines (http://www.ismpp.org/gpp3).

## Contributions

NR, SD, ML, CT, FB conceived the study; CP, GG, PB, MC conceived the study and performed the serological assays. FR, SP, MP, JL collected patients’ samples; NC, IC, NR, CT, ML, interpreted the serological assays; all authors contributed to data acquisition, data analysis, and/or data interpretation, and all authors reviewed and approved the final manuscript.

