## Supplemental material for "Performance characteristics of the VIDAS^®^ SARS-COV-2 IgM and IgG serological assays"

### Table S1. Precision of the VIDAS® SARS-CoV-2 IgM and IgG assays

| VIDAS® SARS-CoV-2 assay | Sample | Total number of measurements | Mean index | Repeatability (within-run precision) | | Within-laboratory precision | |
|---|---|---|---|---|---|---|---|
| | | | | SD | CV (%) | SD | CV (%) |
| IgM | A (negative, high) | 60 | 0.95 | 0.08 | 8.5 | 0.09 | 9.4 |
| | B (positive) | 60 | 1.50 | 0.14 | 9.3 | 0.16 | 10.7 |
| | C (positive) | 60 | 5.78 | 0.42 | 7.3 | 0.46 | 8.0 |
| IgG | D (negative, high) | 60 | 0.88 | 0.03 | 3.8 | 0.04 | 4.5 |
| | E (positive) | 60 | 1.42 | 0.08 | 5.5 | 0.09 | 6.5 |
| | F (positive) | 60 | 8.04 | 0.48 | 5.9 | 0.55 | 6.9 |

Abbreviations: SD, standard deviation; CV, coefficient of variation

### Table S2. Demographics of pre-pandemic healthy volunteers used for the determination of VIDAS® SARS-CoV-2 IgM and IgG specificity

| VIDAS® SARS-CoV-2 serology testing | IgM | IgG | Combined IgM/IgG |
|---|---|---|---|
| Study population, N (%) | 308 (100%) | 989 (100%) | 308 (100%) |
| Age in years, median (range) | 45 (18-86) | 44 (18-86) | 45 (18-86) |
| Gender, N (%) | | | |
| Male | 143 (46.4%) | 347 (35.1%) | 143 (46.4%) |
| Female | 165 (53.6%) | 642 (64.9%) | 165 (53.6%) |
| Pre-pandemic healthy volunteers, N (%) | | | |
| France | 161 (52.3%) | 842 (85.1%) | 161 (52.3%) |
| United States | 147 (47.7%) | 147 (14.9%) | 147 (47.7%) |

### Table S3. Specificity of the VIDAS® SARS-CoV-2 IgM and IgG assays

| VIDAS® SARS-CoV-2 serology testing | No. samples | No. negative results | Specificity | 95% confidence interval (95% CI) |
|---|---|---|---|---|
| **IgM** | 308 | 306 | 99.4% | 97.7-99.9% |
| **IgG**[a] | 989 | 988 | 99.9% | 99.4-100.0% |
| **Combined IgM/IgG**[b] | 308 | 306 | 99.4% | 97.7-99.9% |

[a]Specificity (95% CI) determined based on common samples (N=308) was 100.0% (98.8-100.0%); [b]combined test is negative when both IgM and IgG tests are negative

**Table S4. Positive percent agreement (PPA) of the VIDAS® SARS-CoV-2 IgM and IgG test results according to the time from RT-PCR-positive test result (all available samples)**

| VIDAS® SARS-CoV-2 serology testing | Time from RT-PCR-positive result | Median (range) time in days | No. samples[a] | No. positive results | PPA | 95% CI |
|---|---|---|---|---|---|---|
| IgM (N=234)[b] | 0-7 days | 1.5 (0-7) | 112 | 51 | 45.5% | 36.6-54.8% |
| | 8-15 days | 14 (8-15) | 60 | 49 | 81.7% | 70.1-89.4% |
| | 16-23 days | 20 (16-23) | 38 | 31 | 81.6% | 66.6-90.8% |
| | 24-31 days | 26 (24-28) | 13 | 13 | 100.0% | 75.3-100.0% |
| | ≥ 32 days | 33 (32-65) | 11 | 9 | 81.8% | 52.3-94.9% |
| IgG (N=253)[b] | 0-7 days | 1 (0-7) | 115 | 47 | 40.9% | 32.3-50.0% |
| | 8-15 days | 14 (8-15) | 73 | 63 | 86.3% | 76.6-92.4% |
| | 16-23 days | 20 (16-23) | 41 | 39 | 95.1% | 83.5-99.4% |
| | 24-31 days | 26 (24-28) | 13 | 13 | 100.0% | 75.3-100.0% |
| | ≥ 32 days | 33 (32-65) | 11 | 11 | 100.0% | 71.5-100.0% |

[a]no more than one test result per patient per time period; [b]PPA (95% CI) for common samples (N=232) are shown in Table 4

**Table S5. Positive percent agreement (PPA) of the VIDAS® SARS-CoV-2 IgM and IgG test results according to the time from symptom onset (all available samples)**

| VIDAS® SARS-CoV-2 serology testing | Time from symptom onset | Median (range) time in days | No. samples[a] | No. positive results | PPA | 95% CI |
|---|---|---|---|---|---|---|
| IgM (N=108)[b] | 0-7 days | 6 (1-7) | 24 | 9 | 37.5% | 21.2-57.3% |
| | 8-15 days | 12 (8-15) | 29 | 24 | 82.8% | 65.5-92.4% |
| | 16-23 days | 18 (16-23) | 26 | 26 | 100.0% | 86.8-100.0% |
| | 24-31 days | 26 (24-30) | 18 | 18 | 100.0% | 81.5-100.0% |
| | ≥ 32 days | 34 (32-65) | 11 | 11 | 100.0% | 71.5-100.0% |
| IgG (N=113)[b] | 0-7 days | 5.5 (1-7) | 24 | 9 | 37.5% | 21.2-57.3% |
| | 8-15 days | 12 (8-15) | 31 | 25 | 80.6% | 63.7-90.8% |
| | 16-23 days | 18 (16-23) | 29 | 28 | 96.6% | 82.2-99.9% |
| | 24-31 days | 26 (24-30) | 19 | 18 | 94.7% | 75.4-99.1% |
| | ≥ 32 days | 35 (32-65) | 10 | 10 | 100.0% | 69.2-100.0% |

[a]no more than one test result per patient per time period; [b]PPA (95% CI) for common samples (N=105) are shown in Table 5

**Table S6. Positive percent agreement (PPA) of the VIDAS® SARS-CoV-2 combined IgM/IgG test results and respective number of IgM and IgG positive test results of SARS-CoV-2-positive samples, according to the time from RT-PCR-positive result and of symptom onset**

| VIDAS® SARS-CoV-2 serology testing | Time window in days[a] | Median (range) time in days[a] | No. samples[b] | No. positive results[c] | VIDAS® IgM positive & IgG negative | VIDAS® IgM positive & IgG positive | VIDAS® IgM negative & IgG positive | PPA | 95% CI |
|---|---|---|---|---|---|---|---|---|---|
| **Results relative to documented date of RT-PCR-positive test** | | | | | | | | | |
| **Combined IgM/IgG[c] (N=232)** | 0-7 | 2 (0-7) | 110 | 59 | 9 | 40 | 10 | 53.6% | 44.4-62.7% |
| | 8-15 | 14 (8-15) | 60 | 57 | 4 | 45 | 8 | 95.0% | 86.1-99.0% |
| | 16-23 | 20 (16-23) | 38 | 38 | 2 | 29 | 7 | 100.0% | 90.7-100.0% |
| | 24-31 | 26 (24-28) | 13 | 13 | 0 | 13 | 0 | 100.0% | 75.3-100.0% |
| | ≥ 32 | 33 (32-65) | 11 | 11 | 0 | 9 | 2 | 100.0% | 71.5-100.0% |
| **Results relative to documented date of symptom onset** | | | | | | | | | |
| **Combined IgM/IgG[c] (N=105)** | 0-7 | 5.5 (1-7) | 22 | 8 | 1 | 6 | 1 | 36.4% | 19.7-57.0% |
| | 8-15 | 12 (8-15) | 29 | 26 | 1 | 23 | 2 | 89.7% | 73.6-96.4% |
| | 16-23 | 18 (16-23) | 26 | 26 | 1 | 25 | 0 | 100.0% | 86.8-100.0% |
| | 24-31 | 26 (24-30) | 18 | 18 | 1 | 17 | 0 | 100.0% | 81.5-100.0% |
| | ≥ 32 | 35 (32-65) | 10 | 10 | 0 | 10 | 0 | 100.0% | 69.2-100.0% |

[a]Time from RT-PCR-positive test result (upper Table) or from symptom onset (lower Table); [b]no more than one patient’s test result per time period; [c]combined test is positive when at least one of IgM and/or IgG tests are positive

**Table S7. Positive and negative predictive values (PPV/NPV) at 5% prevalence of the VIDAS® SARS-CoV-2 IgM, IgG and combined IgM/IgG test results, according to the time from symptom onset (all available samples for IgG specificity)**

| VIDAS® SARS-CoV-2 serology testing | Time from symptom onset | PPV[a] (95% CI) | NPV[a] (95% CI) |
|---|---|---|---|
| **IgM** | 0-7 days | 72.1% (36.3-92.1%) | 96.5% (95.4-97.4%) |
| | 8-15 days | 87.0% (62.5-96.4%) | 99.1% (98.0-99.6%) |
| | 16-23 days | 89.0% (67.1-97.0%) | 100.0% (N/A)[b] |
| | 24-31 days | 89.0% (67.1-97.0%) | 100.0% (N/A)[b] |
| | ≥ 32 days | 89.0% (67.1-97.0%) | 100.0% (N/A)[b] |
| **IgG** | 0-7 days | 94.3% (68-99.2%) | 96.5% (95.4-97.4%) |
| | 8-15 days | 97.8% (86.3-99.7%) | 99.3% (98.2-99.7%) |
| | 16-23 days | 98.0% (87.6-99.7%) | 99.8% (98.6-100.0%) |
| | 24-31 days | 98.0% (87.4-99.7%) | 99.7% (98.1-100.0%) |
| | ≥ 32 days | 98.1% (88-99.7%) | 100.0% (N/A)[b] |
| **Combined IgM/IgG** | 0-7 days | 74.7% (40-92.9%) | 96.7% (95.6-97.6%) |
| | 8-15 days | 87.9% (64.5-96.7%) | 99.5% (98.4-99.8%) |
| | 16-23 days | 89.0% (67.1-97.0%) | 100.0% (N/A)[b] |
| | 24-31 days | 89.0% (67.1-97.0%) | 100.0% (N/A)[b] |
| | ≥ 32 days | 89.0% (67.1-97.0%) | 100.0% (N/A)[b] |

[a]PPV and NPV were calculated at 5% prevalence using values of sensitivity (PPA) determined on paired IgM and IgG test results (N=105; Table 5); values of specificity were those calculated on all available samples (N=308 for IgM and N=989 for IgG; Table 3); [b]95% CI not calculable (division by zero)
